# Role of type I hypersensitivity reaction in the development of overall and uncomplicated acute appendicitis: a systematic review and meta-analysis

**DOI:** 10.1101/2024.06.11.24308763

**Authors:** Javier Arredondo Montero, María Rico Jiménez, Blanca Paola Pérez Riveros, Rafael Fernández Atuan, Niklas Pakkasjärvi, Nellai Krishnan, Carlos Delgado-Miguel, Sachit Anand

## Abstract

**Background:** This systematic review aimed to analyze the potential etiopathogenic role of a type I hypersensitivity reaction in the development of overall acute appendicitis (AA), non-complicated acute appendicitis (NCAA), and complicated acute appendicitis (CAA).

**Methods:** This review was prospectively registered in PROSPERO (CRD42024516547). We included both prospective and retrospective original clinical studies that examined the role of immunoallergic processes in the development of acute appendicitis (AA). A comprehensive search was conducted in PubMed, Web of Science, Scopus, and OVID, using the following search terms and keywords: (allergy OR allergic OR immunoallergy OR immunoallergic OR immunomediated) AND (appendicitis OR appendectomy) AND (IgE OR “IgE-mediated” OR hypersensitivity OR “type I”). Two independent reviewers meticulously selected the articles and extracted relevant data. The methodological quality of the studies was rigorously assessed using the Newcastle-Ottawa index. A synthesis of the results, a standardization of the metrics, and seven random-effect meta-analyses were performed.

**Results:** This review included nineteen studies. A random-effects meta-analysis including six articles (6370 patients with NCAA and 2000 patients with CAA) showed that patients with any documented history of IgE-mediated allergy had a lower risk of developing CAA (OR 0.52, 95%CI [0.38-0.72], p<0.0001). The random-effect meta-analysis for serum Interleukin-9 (NCAA vs. CAA) included two articles (177 patients with NCAA and 101 patients with CAA) and resulted in a significant mean difference [95% CI] of –0.38 [-0.67,-0.08] pg/mL (p=0.01). The random-effect meta-analysis for serum Interleukin-13 (NCAA vs. CAA) included two articles (177 patients with NCAA and 101 patients with CAA) and resulted in a significant mean difference [95% CI] of –11.32 [-13.90,-8.75] pg/mL (p=<0.00001). The random-effect meta-analysis for total eosinophil count (NCAA vs. CAA) included three articles (455 NCAA and 303 CAA) and resulted in a significant mean difference [95% CI] of –0.06 [-0.09,-0.04] eosinophils x 10^9^/L (p=<0.00001).

**Conclusions:** The present study demonstrates an association between a type I hypersensitivity reaction and the development of NCAA. Additionally, our meta-analytic model shows significantly higher levels of eosinophils peripheral blood in patients with NCAA than in patients with CAA. These findings suggest a potential immunoallergic mediation in the development of NCAA. Future prospective studies must validate these findings since these patients may benefit from specific therapeutic targets.

**Funding:** None

**Registration:** PROSPERO (CRD42024516547).

## Introduction

Acute appendicitis (AA) is the most frequent urgent surgical pathology in the world, with a lifetime risk of 6 to 8% [1]. In recent years, significant advances have been made in therapeutic terms in this pathology, such as the safe introduction of minimally invasive techniques [2]. Nevertheless, this continues to be a pathology with a crucial diagnostic error rate, and there is an essential lack of knowledge regarding its etiopathogenesis. In recent years, multiple biomarkers have been evaluated as potential diagnostic tools in AA. A prominent example is serum interleukin-6 (IL-6), which has demonstrated excellent diagnostic capability for diagnosing AA and an excellent ability to discriminate between complicated appendicitis (CAA) and non-complicated appendicitis (NCAA) [3]. Other systemic inflammatory biomarkers, such as pentraxin-3 [4], have also been evaluated, with variable diagnostic yields. However, the immune and inflammatory response associated with AA is now considered complex and involves multiple distinct signaling pathways.

In recent years, increasing evidence has emerged for the potential role of type I hypersensitivity reactions in the development of AA. These reactions follow antigen presentation to naive lymphocytes (Th0) via antigen-presenting cells (APCs) and are mediated by multiple cytokines, including Interleukin 4 (IL-4), Interleukin-5 (IL-5), Interleukin-9 (IL-9), and Interleukin-13 (IL-13). These reactions mediate eosinophilic activation, release of eosinophil granule proteins, and mast cell activation with subsequent histamine and tryptase release. [5]. This, in turn, leads to smooth muscle contraction mediated by leukotrienes and histamine, vasodilatation (at the expense of prostaglandins and histamine), and release of proteases with potential tissue damage. All these mechanisms may be potentially involved in the development of AA (i.e., proteases may be related to the occurrence of appendiceal perforation) [6]. Likewise, the appendix is a structure rich in enterochromaffin cells (especially at the level of the lamina propria). These cells store and secrete serotonin, a neurotransmitter that modulates peripheral immune system responses [7,8]. Lastly, the cecal appendix contains a significant representation of lymphoid tissue. Because of these two aspects, the reaction to allergen exposure may be more accentuated in this organ than in the rest of the body [7,8]. This review aims to compile and meta-analyze the existing evidence on the role of type I hypersensitivity reactions in the development of AA.

## Methods

### Literature search and selection

We followed the Preferred Reporting Items for Systematic Reviews and Meta-Analyses (PRISMA) guidance [9]. We specifically designed and implemented a review protocol registered in the International Prospective Register of Systematic Reviews (PROSPERO ID CRD42024516547). Eligible studies were identified by searching the main existing medical bibliography databases (PubMed, Web of Science, Scopus, OVID). The search covered the period from 2008 to 03.2024. Search terms and keywords were (allergy OR allergic OR immunoallergy OR immunoallergic OR immunomediated) AND (appendicitis OR appendectomy) AND (IgE OR “IgE-mediated” OR hypersensitivity OR “type I”). In addition, the bibliographical references of the included papers were screened for potential inclusion in this review. The search was last executed on 03.2024.

Supplementary File 1 shows inclusion and exclusion criteria. JAM and MRJ selected articles using the COVIDENCE ® tool. The search results were imported into the platform, and both authors screened the articles separately. Disagreements were resolved by consensus.

### Quality assessment

The selected articlés methodological quality and risk of bias were evaluated with the Newcastle-Ottawa scale (NOS) [10]. Depending on the study design evaluated, case-control and cohort templates were employed. Selection, comparability, and exposure items were assessed for each included study.

### Data extraction and synthesis

Three independent reviewers (JAM, BPR, MRJ) extracted the relevant data from the selected articles following a standardized procedure. Extracted data included the author, the country where the study was conducted, year of publication, study design, study population (sample size, age range, and sex distribution), AA group and control group (CG) definitions, reference standard used in AA group, number of events (allergy) in each group, mean and standard deviation (or median and interquartile range) for each analyzed biomarker, counts of the different cell subpopulations per unit of histological area/surface area and the statistical p-value for each between-group comparison. There were no disagreements between the reviewers after collating the extracted data. The metrics used in each study were reviewed, and a standardization of units was performed. Medians (Interquartile ranges) were converted to means (standard deviations) following a standardized procedure [11].

### Meta-analysis

A first meta-analytical model was performed for the dichotomous variable “IgE-mediated allergy” and the risk of developing complicated acute appendicitis (CAA). The Mantel-Haenszel method was used. The results were expressed as an odds ratio (95% CI) and were plotted in a forest plot. Between-study heterogeneity was assessed using the Chi^2^ and I^2^ statistics. A funnel plot analysis was performed to evaluate a potential publication bias.

Six additional random-effects meta-analyses were performed: 1) Serum Interleukin-4 (IL-4) (NCAA vs. CAA), 2) Serum Interleukin-5 (IL-5) (NCAA vs. CAA), 3) Serum Interleukin-9 (IL-9) (NCAA vs. CAA) 4) Serum Interleukin-13 (IL-13) (NCAA vs. CAA), 5) Total eosinophil count (TEC) (AA vs. CG) and 6) TEC (NCAA vs. CAA). The results were expressed as a mean difference (95% CI) and were plotted in a forest plot. Between-study heterogeneity was assessed using the Chi^2^ and I^2^ statistics.

## Results

The search returned 203 articles (Scopus n=15; Pubmed n=113; Web of Science n=14; OVID n=61). Thirty-five duplicates were removed. Among the remaining 168 articles, we excluded 148 (inclusion and exclusion criteria, n=148). We excluded one additional work when assessing eligibility due to a wrong study design (n=1). This review finally included 19 studies with data from approximately 121,742 participants [12–30]. Figure 1 shows the flowchart of the search and selection process.

**Figure 1.**
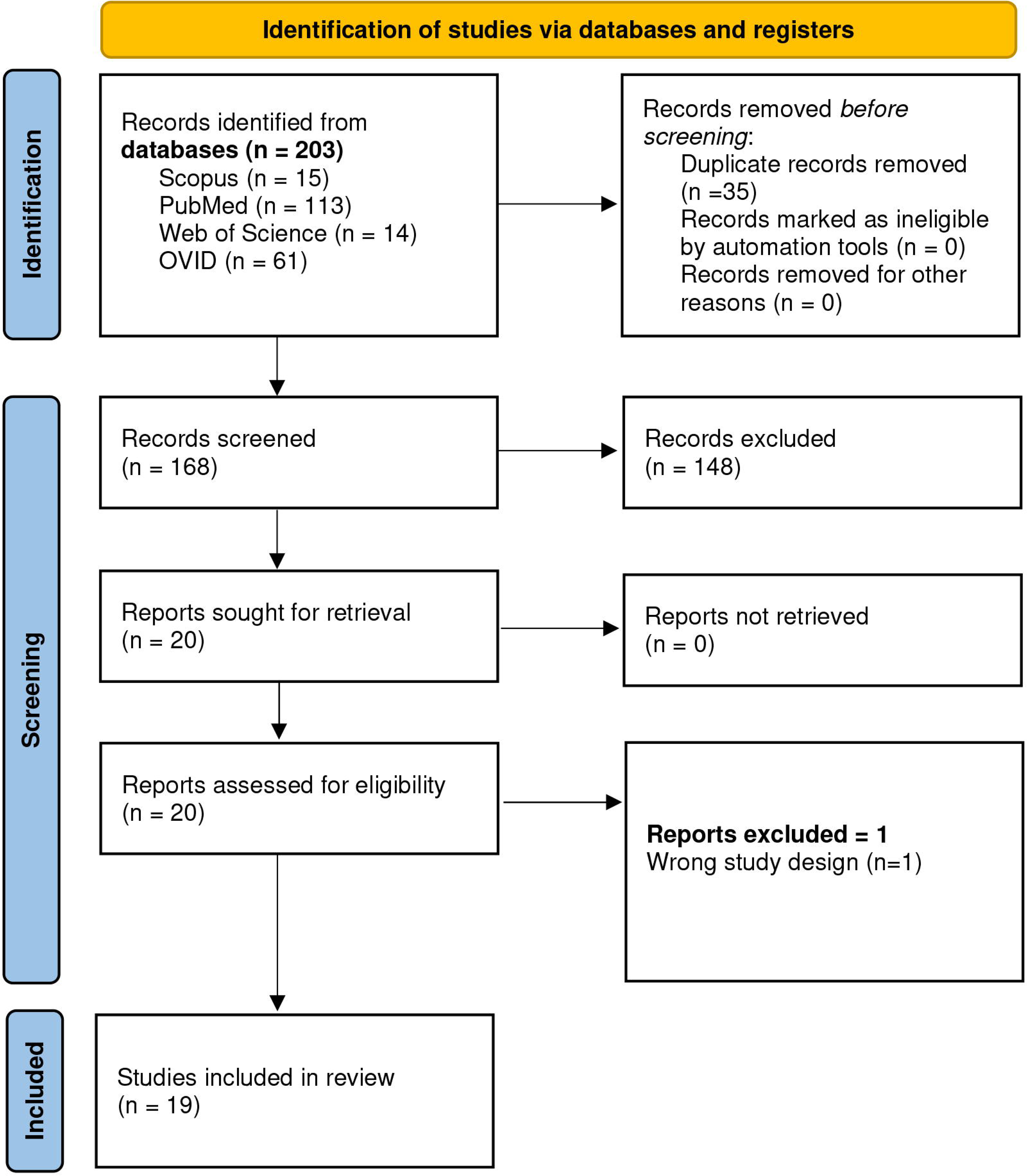
Flowchart of the search and selection process.

The quality of twelve of the studies was considered good [12–18,20,21,23,27,30], and the quality of seven of them was considered fair [19,22,24,25,26,27,29] at the expense of the category “Ascertainment of exposure.” No study was classified as “poor”.

### IgE-mediated allergy and the development of non-complicated acute appendicitis

#### Sociodemographic characteristics

Data from the ten studies that evaluated the role of allergy as a potential protective factor in the development of CAA are shown in Table 1. All analyses were carried out between 2008 and 2024. Two were from Turkey [12,16], three from Sweden [18,23,25], two from Portugal [22,29], one from Korea [20], one from China [27] and one from the USA [30]. Five studies were retrospective [18,20,23,27,30], four were prospective [12,22,25,29], and one was categorized as prospective based on the information contained in the manuscript. However, the authors did not make this explicit [16]. Six studies involved pediatric populations [16,18,23,25,27,30].

The definition of “case” was consistent in five of the selected studies, given as the histopathological confirmation of AA in the surgical specimen [12,16,22,25,29]. Two studies defined “case” as an intraoperative diagnosis of acute appendicitis, although the authors confirmed that they excluded patients with negative appendectomy findings after histopathologic analysis [18,27]. One study defined “case” as an appendectomy based on surgical codes from a national database [20]. One study defined “case” as an “acute appendicitis” episode based on clinical ICD-10 codes from a national database and included non-appendectomized patients in its analysis [23]. One study considered “definite appendicitis” as the confluence of a clinical diagnosis of appendicitis, intraoperative evidence of appendicitis, and histologic confirmation of appendicitis [30]. This was not the case for the definition of “control,” which constituted either sex-matched healthy individuals or ASA 1 patients who were admitted to the Department of General Surgery for other diseases [12], patients attending the pediatric surgery unit for different pathologies, such as circumcision or hernia [16], matched-controls from a national database [20,30], incidental appendectomies [22,29], negative appendectomies [22] and children with abdominal pain due to other diagnoses [25].

Two studies specifically evaluated potential conditions that could have altered allergy tests, including concomitant pathology and recent use of H_2_-receptor antagonists, some tricyclic antidepressants (such as desipramine and doxepin), and antihistaminics [12,16]. The rest of the studies did not specify specific exclusions regarding possible confounders of IgE-mediated allergies.

Two studies assessed the presence of allergies by Mixed Inhalant Prick Test (MPT) [12,16], Food Allergy Test [12,16], or a mixture of both [12,16]. One study relied on retrospective patient records to identify IgE-mediated allergies to aeroallergens, food allergens, and antibiotics [18]. Four studies considered allergy as a medical history allergy reflected in patient records [22,25,27,29]. Three studies relied on national databases for analyses [20,23,30]. Of these three studies, one study compared a national database of appendectomized and non-appendectomized patients and compared the prevalence of asthma in both groups [20]. Another used a national database and followed a previously established protocol to identify the allergy diagnosis based on ICD-10 codes and codes of allergy-specific medications previously dispensed or prescribed [23]. Lastly, another study compared the prevalence of active asthma between the AA group and the CG [30].

All ten studies reported the data as the number of allergy positives out of the total number of patients per group [12,16,18,20,22,23,25,27,29,30].

#### Random-effects meta-analysis for IgE-mediated allergy and the development of NCAA

A random-effects meta-analysis including six articles (6370 patients with NCAA and 2000 patients with CAA) showed that patients with any documented history of IgE-mediated allergy had a lower risk of developing CAA (OR 0.52, 95%CI [0.38-0.72], p<0.0001). The I^2^ value was 49%, and the Chi^2^ was 9.72. The graphical depiction of this analysis is shown in Figure 2. The graphical depiction of the funnel plot associated with this analysis is shown in Figure 3. It was considered symmetric (no publication bias).

**Figure 2.**
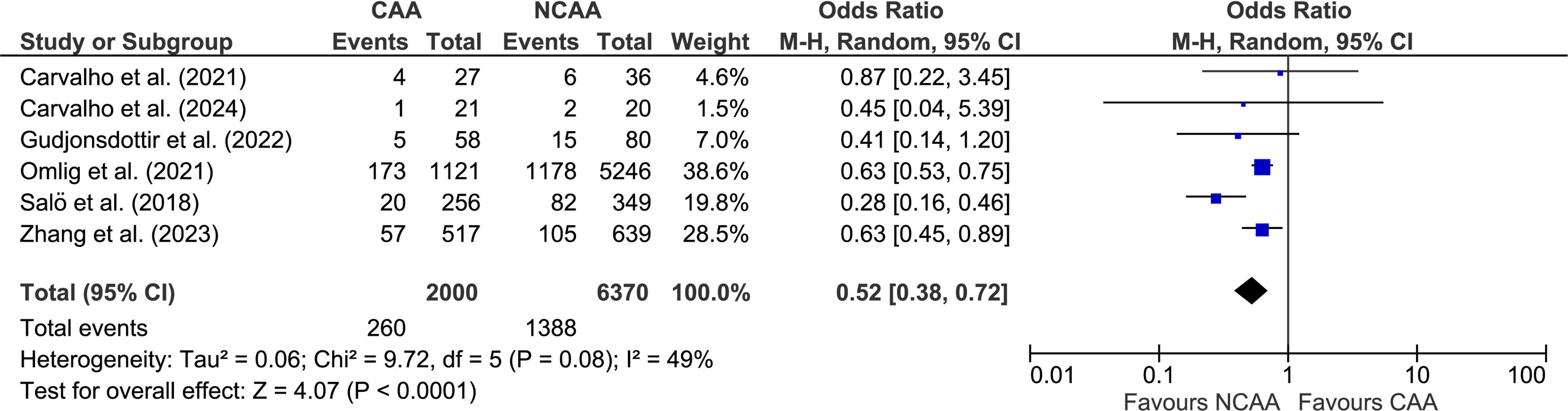
Forest plot of the random-effects meta-analysis evaluating the relationship between IgE-mediated allergy and the development of CAA (Mantel-Haenszel method).

**Figure 3.**
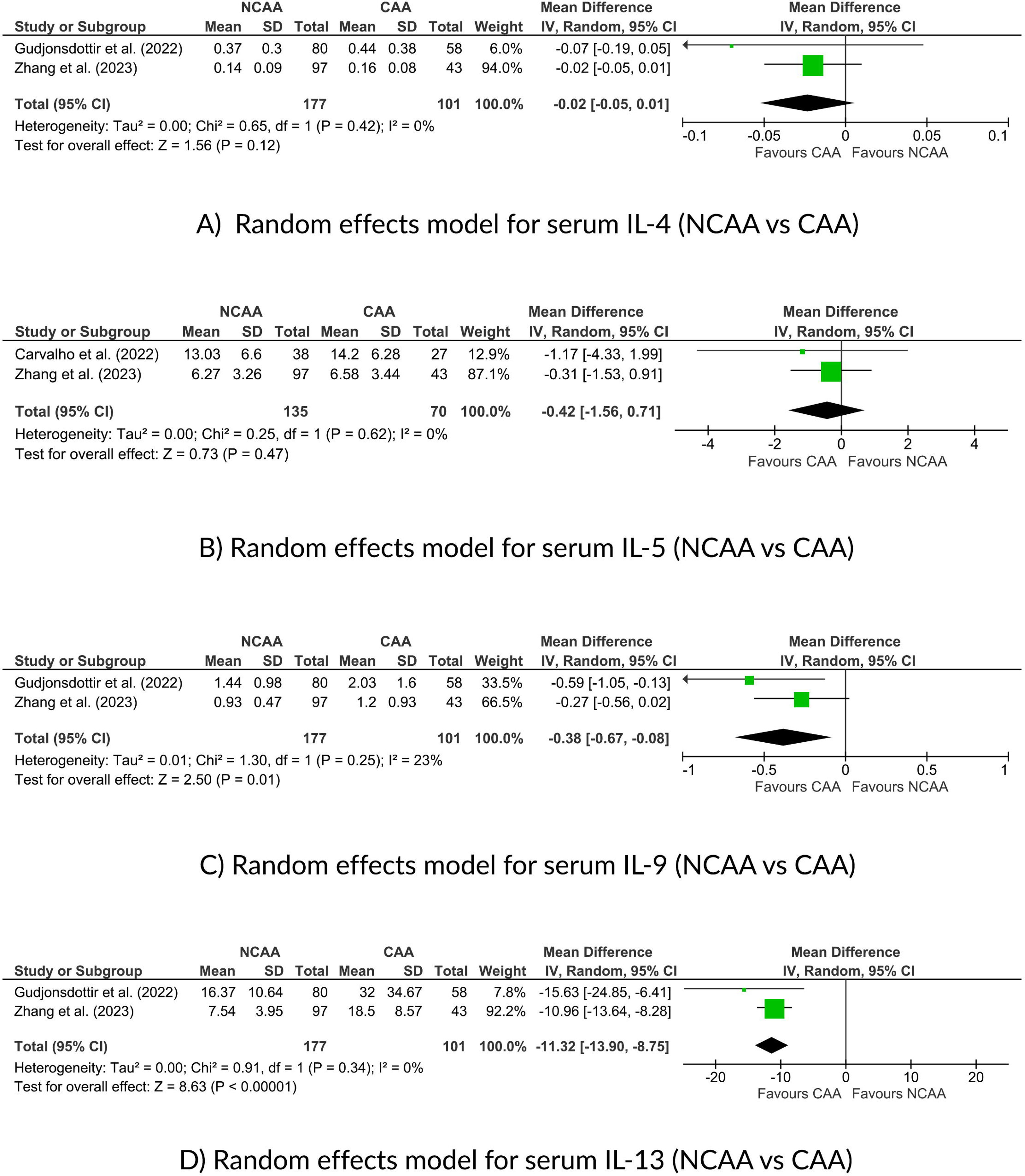
Funnel plot concerning the random-effects meta-analysis evaluating the relationship between IgE-mediated allergy and the development of CAA.

### Type I hypersensitivity reaction-related biomarkers and acute appendicitis

#### Sociodemographic characteristics

Table 2 shows data from the six studies that evaluated the role of different allergy-related biomarkers in acute appendicitis [19,22,24,25,28,29]. All studies were carried out between 2019 and 2024. Four were from Portugal [19,22,24,29], one from Sweden [25], and one from China [28]. All studies were prospective [19,22,24,25,28,29]. Two studies involved pediatric populations [25,28].

The definition of “case” was consistent in all the selected studies, given as the histopathological confirmation of AA in the surgical specimen [19,22,24,25,28,29]. This was not the case for the definition of “control,” which was constituted by incidental appendectomies [22,24,29], normal histologic findings in the cecal appendix [19,22,24], and children with abdominal pain due to other diagnoses [25]. We believe that using a control group with colorectal carcinoma should be considered a limitation of the work of Carvalho et al. [29]. Although we chose not to exclude this work from the review, we believe that this may have conditioned the results of the analyses. Zhang et al. [28] did not have a control group.

Appendicular lavage fluid (ALF) samples [19,24,29], immunohistochemical techniques on the appendix tissue [22], and serum interleukin and IgE measurements [24,25,28] were assessed.

Two studies expressed their results as mean (standard deviation) [19,28], and four studies expressed their results as median (interquartile range) [22,24,25,29]. Zhang et al. [28] expressed serum IL-6 values as median (interquartile range). When it was considered necessary for developing meta-analytical models, an inferential estimation of the mean and standard deviation based on the median and interquartile range was performed. Both the original and inferred values are shown in Table 2.

Concerning units, serum interleukins were expressed in pg/mL and IgE in ng/mL. No unit conversion was required.

#### Appendicular lavage fluid biomarkers

Three papers belonging to the same group of authors evaluated different biomarkers in ALF [19,24,29]. The first work [19] showed statistically significant differences in IL4 and IL-5 levels between the three groups included in the study (Kruskal-Wallis test, three-group comparison). In the second study [24] obtained a marginal significance for IL-5 in similar conditions and with similar groups. In the third study, they evaluated the levels of histamine, serotonin, and tryptase in ALF, finding statistically significant differences between the three groups for the levels of tryptase (p=0.001) [29].

#### Immunohistochemical IgE-positive cells assessment

Carvalho et al. [22] demonstrated statistically significant differences in the number of IgE-positive cells when comparing NCAA and CG.

#### Serum Interleukins and serum IgE

Serum levels of IL-4 [25,28], IL-5 [24,28], IL-6 [28], IL-9 [25,28] and IL-13 [25,28] were assessed. Statistically significant differences between the NCAA and CAA groups were found for IL-6 [28], IL-9 [25,28], and IL-13 [25,28].

Gudjonsdottir et al. [25] found no statistically significant differences in serum IgE levels between the NCAA and CAA groups or between the AA and control groups. In contrast, Zhang et al. demonstrated statistically significant differences in serum IgE levels between the NCAA and CAA groups (p<0.01) [28].

#### Random-effects meta-analysis for serum interleukins (NCAA vs. CAA)

Four random-effects meta-analyses were performed: 1) Serum Interleukin-4 (IL-4) (NCAA vs. CAA), 2) Serum Interleukin-5 (IL-5) (NCAA vs. CAA), 3) Serum Interleukin-9 (IL-9) (NCAA vs. CAA) 4) Serum Interleukin-13 (IL-13) (NCAA vs. CAA). The random-effect meta-analysis for serum Interleukin-9 (NCAA vs. CAA) included two articles (177 patients with NCAA and 101 patients with CAA) and resulted in a significant mean difference [95% CI] of –0.38 [–0.67, –0.08] pg/mL (p=0.01). The I^2^ value was 23%, and the Chi^2^ was 1.30. The random-effect meta-analysis for serum Interleukin-13 (NCAA vs. CAA) included two articles (177 patients with NCAA and 101 patients with CAA) and resulted in a significant mean difference [95% CI] of – 11.32 [-13.90,-8.75] pg/mL (p=<0.00001). The I^2^ value was 0%, and the Chi^2^ was 0.91. Results are shown in Figure 4.

**Figure 4.**
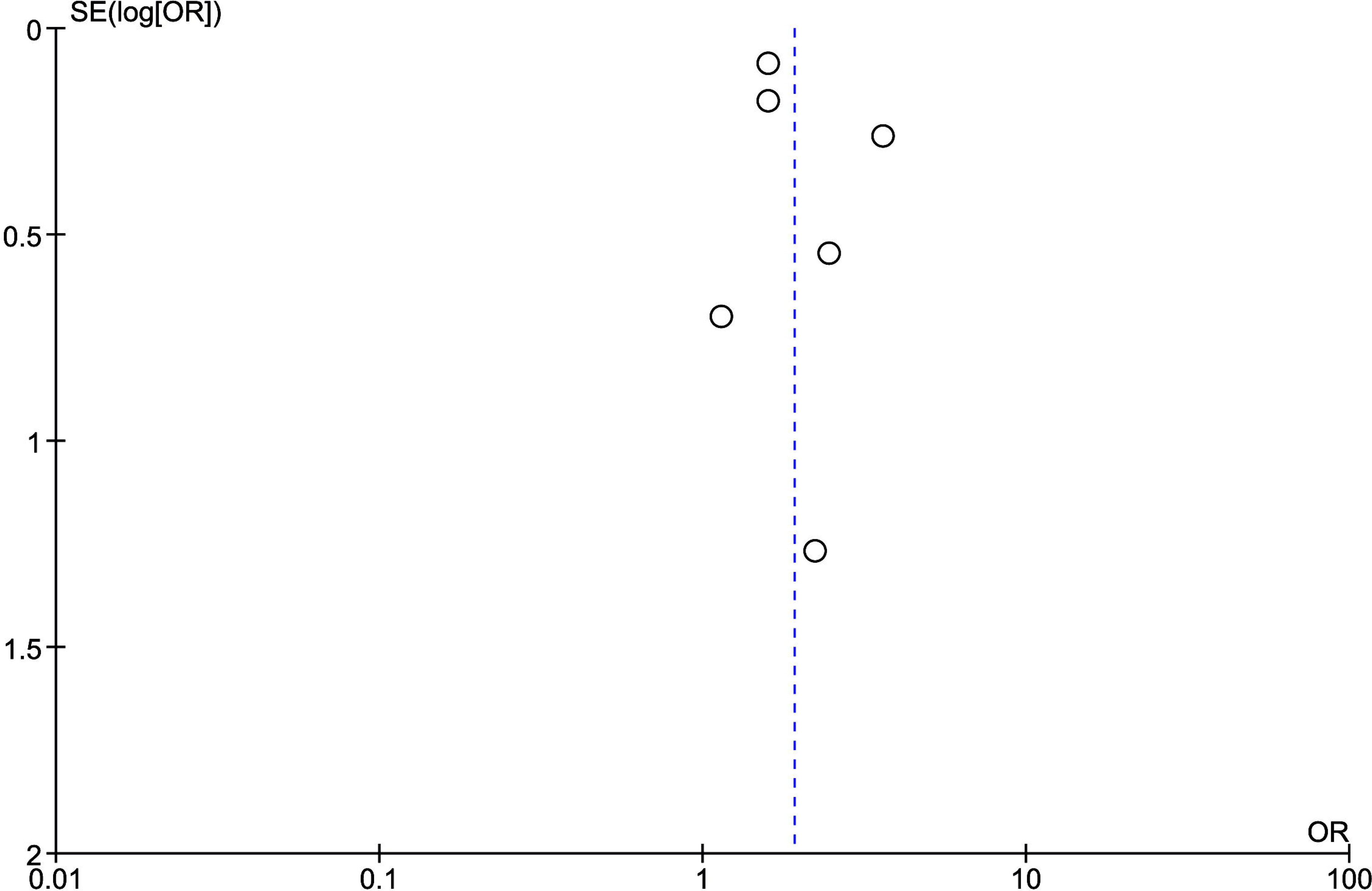
Forest plot of the random-effects meta-analysis for serum interleukins (NCAA vs. CAA) (Inverse variance method). A) serum IL-4; <u>B</u>) serum IL-5; <u>C</u>) serum IL-9; <u>D</u>) serum IL-13.

### Eosinophils, Mast cells, and acute appendicitis

#### Sociodemographic characteristics

Table 3 shows data from the nine studies that evaluated the role of eosinophils and mast cells in acute appendicitis. All studies were carried out between 2008 and 2023. Three were from India [13,14,21], three from Turkey [12,15,16], two from Portugal [24,26], and one from China [17]. Five studies were prospective [12,13,21,24,26]. Two studies were retrospective [14,17], and one was categorized as prospective based on the information contained in the manuscript. However, the authors did not make this explicit [16]. One study did not provide information concerning study design [15]. Three studies involved a pediatric population [15–17].

The definition of “case” was consistent in all the selected studies, given as the histopathological confirmation of AA in the surgical specimen [12–17,21,24,26]. This was not the case for the definition of “control,” which was constituted either by histologically normal appendectomy specimens [13–15,21,24,26], healthy volunteers [13], patients with acute bronchial asthma [13], patients attending the pediatric surgery unit for other pathologies, such as circumcision or hernia [16] or sex-matched healthy individuals or ASA 1 patients who were admitted to the Department of General Surgery for different diseases [12]. Minderjahn et al. did not report a control group [17].

Four studies expressed their results as mean (standard deviation) [12,14,16,17], one study as median (range) [13], one study as mean [15], one study as mean (95% confidence interval) [21], and two studies as median (interquartile range) [24,26]. When it was considered necessary for developing meta-analytical models, an inferential estimation of the mean and standard deviation based on the median and interquartile range was performed. Both the original and inferred values are shown in Table 3.

Concerning units, TEC in peripheral blood, results were expressed in cells/x10^9/L [17,24,26] or in cells/mm^3^ [12]. Although most authors expressed TEC units correctly, in the case of Cevizci et al. [16], TEC units were not listed in the paper. We tried to contact the authors to clarify this point but failed. To develop the meta-analytic model, we converted it to an order of magnitude with the highest biological plausibility and compatibility with the other reported studies. Since peripheral blood eosinophils follow a relatively narrow distribution range in humans, we believe this was the correct approach. Still, it must be considered a limitation due to its inferential nature. Similarly, in the case of Harlak et al. [12], the units reported were in cells/mm^3^, so we performed the conversion to International Units as shown in Table 3.

Eosinophil cationic protein, eosinophil-derived neurotoxin, and eosinophil peroxidase were expressed as ng/mL (or as µg/L, which has a conversion ratio of 1:1) both in serum and in ALF [13,26]. The presence of eosinophils, mast cells, and Cajal cells at the histological level was performed by counting per unit area or field with variable methodologies [12,13,14,15,21,24].

#### Histologic assessment of Cajal cells and acute appendicitis

Karakuş et al. reported statistically significant differences in the density mean values of Cajal cells when comparing the control group and the AA group (p<0.05) [15].

#### Histologic assessment of mast cells and acute appendicitis

Two studies [14,21] evaluated by immunohistochemical techniques (CD117) the presence of mast cells in quantitative terms. Mohan et al. reported statistically significant differences in the number of degranulated mast cells in submucosa per whole cross-sectional area (p=0.01). Karakuş et al. reported statistically significant differences in the density mean values of Mast cells when comparing the control group and the AA group (p<0.05) [15].

#### Histologic assessment of eosinophils and acute appendicitis

Six studies evaluated the degree of eosinophilic infiltration in the cecal appendix at the histological level [12,13,14,15,21,24]. Santhosh et al. reported statistically significant differences in the mean eosinophil count in the appendix (number of eosinophils per high power field) when comparing AA and the control group [13]. Mohan et al. [21] also reported statistically significant differences between groups in the eosinophil count in the muscle layer of the appendix per unit area. Interestingly, these differences became more noticeable when a whole cross-sectional area was evaluated. Carvalho et al. [24] also reported statistically significant differences in the eosinophilic infiltration rate of the appendiceal wall between the three groups being assessed in the study (p<0.001). Karakuş et al. reported statistically significant differences in the density mean values of eosinophils when comparing the control group and the AA group (p<0.05) [15].

#### Peripheral blood eosinophils and acute appendicitis

Two studies evaluated serum eosinophil cationic protein (SECP) [13,26]. Both found statistically significant differences between AA and controls.

Five studies evaluated TEC in peripheral blood [12,16,17,24,26]. Minderjahn et al. reported statistically significant differences between the NCAA and CAA groups (p<0.0001) [17]. Carvalho et al. reported statistically significant differences between the three groups (control group, NCAA, and CAA) in one of their works [24] and marginal significance for another of their analyses in similar conditions [26].

#### Random-effects meta-analysis for total eosinophil count (peripheral blood)

Two random effects models were performed: 1) TEC (AA vs. CG) and 6) TEC (NCAA vs CAA). The random-effect meta-analysis for total eosinophil count (AA vs. CG) included four articles (287 AA and 256 controls) and resulted in a significant mean difference [95% CI] of 0.01 [0.00, 0.03] eosinophils x 10^9^/L (p=0.006). The I^2^ value was 0%, and the Chi^2^ was 2.18. The random-effect meta-analysis for total eosinophil count (NCAA vs. CAA) included three articles (455 NCAA and 303 CAA) and resulted in a significant mean difference [95% CI] of –0.06 [–0.09, –0.04] eosinophils x 10^9^/L (p=<0.00001). The I^2^ value was 33%, and the Chi^2^ was 2.99. Results are shown in Figure 5.

**Figure 5.**
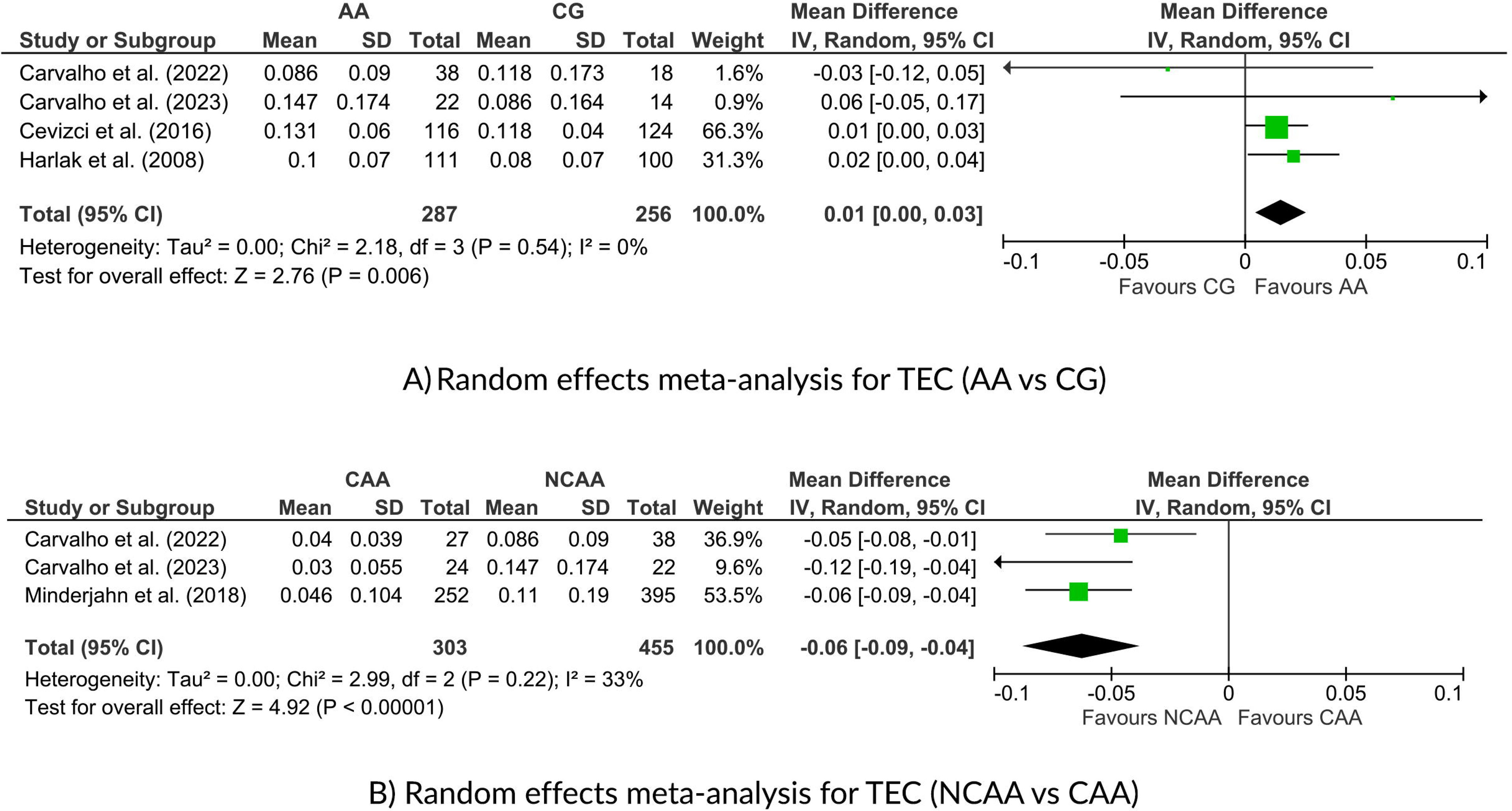
Forest plot of the random-effects meta-analysis for total eosinophil count in peripheral blood (Inverse variance method). <u>A</u>) AA vs. CG; <u>B</u>) NCAA vs. CAA.

## Discussion

In the present work, we systematically reviewed the role of type I hypersensitivity reactions in the development AA and NCAA. Through performing seven meta-analytical models, several key findings emerged: 1) the presence of type I hypersensitivity reactions (IgE-mediated allergy) is a protective factor for the development of CAA; 2) there are significantly higher levels of serum IL-9 and IL-13 in patients with CAA than in patients with NCAA; 3) the total eosinophil count in peripheral blood is significantly higher in patients with AA than in controls and is significantly higher in NCAA than in CAA.

The reviewed literature consistently demonstrates a relationship between IgE-mediated allergy and the development of NCAA. Case-control and cohort studies show a higher prevalence of IgE-mediated allergy in patients with AA, specifically in patients with NCAA. Additionally, national cohort studies show a higher rate of appendectomies in patients with asthma and a higher rate of asthma in patients with AA, providing additional support for this theory.

Concerning the cytokine profile, IL-9 and IL-13 belong to the cytokine signaling chain of type I hypersensitivity reactions. Again, its elevation in AA cases compared to controls supports the presence of a type I hypersensitivity reaction in AA. Although following this concept, it would be expected to see higher levels in the NCAA group, their significantly higher levels in the CAA group are attributable, in our opinion, to the more evolved process and more remarkable systemic inflammation. It should also be considered that the synthesis, production, and release of these cytokines are not immediate and require several hours from the onset of the process. Previous findings with IL-6 support this theory.

Eosinophils are an earlier marker that, in our opinion, correlates better with a potential type I hypersensitivity reaction occurring at the level of the cecal appendix. In many cases, peripheral eosinophilia may be masked by the marked neutrophilia characteristic of AA, so this may be a subtle finding. Using specific eosinophilic peripheral blood markers, such as serum eosinophil cationic protein, eosinophil peroxidase, or eosinophil-derived neurotoxin, seems a promising line of research for the future. On the other hand, the reviewed literature consistently shows higher tissue-level eosinophil levels in patients with AA than in controls, which supports the etiopathogenic hypothesis put forward in the present manuscript. We have observed that the reporting of eosinophils at the histological level presents a significant methodological heterogeneity in the literature. This field could benefit from artificial intelligence (AI) models in quantitative terms (such as convolutional neural networks).

Based on our findings, we postulate that AA could encompass two distinct nosological entities: on the one hand, a subtype of NCAA, which could have an immunoallergic etiopathogenesis mediated by a type I hypersensitivity reaction, and on the other hand, an NCAA of another etiology (such as bacterial infection, endoluminal obstruction, foreign body, parasites…) that invariably will progress to CAA (i.e. by a decrease in parietal tissue perfusion pressure secondary to endoluminal obstruction leading to gangrene or by action of local metalloproteases with subsequent appendicular perforation). At present, we believe that there is not enough evidence to discriminate which NCAA could have a strictly immunoallergic behavior and be self-limiting and which NCAA (even having an immunoallergic etiology) will progress to CAA, except for those with an obvious underlying cause, such as an endoluminal appendicolith. K. P. Aravindan, the first author to postulate this possible etiopathogenesis for AA, stated that one explanation is that the infection was a later consequence of a type I hypersensitivity reaction [31]. If this were the case, an early rupture of the type I hypersensitivity reaction could constitute an effective and early treatment for AA in the future.

These findings could have profound implications for treating NCAA associated with a type I hypersensitivity reaction if confirmed. The possibility of treating such cases with drugs like antihistamines or corticosteroids would mark a paradigm shift in our understanding and management of the disease. However, before considering this scenario, we must develop prospective multicenter studies to confirm and validate our findings, focusing on improving the discriminatory capacity of the subgroup of patients with an underlying type I hypersensitivity reaction. Furthermore, the high recurrence rates of AA in conservatively managed patients align with this hypothesis, suggesting that the type I hypersensitivity reaction may be reproduced upon new antigenic exposure [32].

The present study presents important strengths, including a robust methodology adhering to the PRISMA guidelines and executing multiple meta-analytical models. The low heterogeneity obtained in some of these models should also be considered an additional strength. However, despite these strengths, this study also faces significant limitations. These include the low sample size observed in some of the included analyses, heterogeneity in the definition of the control groups, and the retrospective nature of certain studies. Furthermore, methodological heterogeneity in reported histological evaluations constitutes a significant limitation.

In conclusion, the evidence suggests a role for type I hypersensitivity reactions in the development of NCAA. Future studies are essential to validate these findings and enable the stratification of patients based on this factor, potentially facilitating the identification of specific therapeutic targets for this subgroup.

## ETHICAL APPROVAL

This study did not involve the participation of human or animal subjects, and therefore, institutional review board approval was not sought.

## STATEMENT OF AVAILABILITY OF THE DATA USED DURING THE SYSTEMATIC REVIEW

The data used to carry out this systematic review are available upon request from the review authors.

## Supporting information

PRISMA checklist

## Data Availability

All data produced in the present study are available upon reasonable request to the authors

