## Supplementary material for "Role of type I hypersensitivity reaction in the development of overall and uncomplicated acute appendicitis: a systematic review and meta-analysis": Table 1

| **Author** | **Country** | **Study design** | **Age (Range)** | **Sex M/F** | **Total *N*** | ***N* in AA** | ***N* in CG** | **Allergy assessment** | **P**  **(AA vs CG)** | **P (NCAA vs CAA)** |
| --- | --- | --- | --- | --- | --- | --- | --- | --- | --- | --- |
| Harlak et al. (2008) [12] | Turkey | Prospective case-control | 14-57y | 171/40 | 211 | 111 | 100 | **MPT**: AA: 33/111, CG: 7/100  **FAT:** AA: 14/111, CG: 1/100  **MPT+FAT**: AA: 38/111, CG: 8/100 | **MPT**:<0.001  **FAT**: <0.001  **MPT + FAT**: < 0.001 | - |
| Cevizci et al. (2016)* [16] | Turkey | Prospective case-control****** | **AA**: 10.1 (2.9)y^1^  **CG**: 9.8 (2.6)y^1^ | 140/100 | 240 | 116 | 124 | **MPT**: **SA**: 1.7%, **PA**: 3.4%, **CAA**: 0%  **FAT**: AA**:** 2.6%, **SA**: 0.9%, **PA**: 3.4%, **CAA**: 0%  **MPT+FAT**: AA**:** 1.7%, **SA**: 0.9%, **PA**: 4.3%, **CAA**: 0% | >0.05 | - |
| Hasassri et al. (2016)* [30] | USA | Retrospective Cohort | <18ª | 370/248 | 618 | 309 | 309 | **Active asthma:**  **AA:** 41/309**, CG:**23/309  **OR:** 1.88 [1.07-3.27]  **Inactive asthma:**  **AA:** 35/309**, CG:** 46/309  **OR**: 0.76 [0.46-1.26] | - | **-** |
| Salö et al. (2018)* [18] | Sweden | Retrospective Cohort | <15y | 381/224 | 605 | 605 **NCAA:** 349**, CAA:** 256 | - | **IgE-mediated allergy:**  **Pollen: NCAA**: 44/349, **CAA**: 7/256  **Fur/mite: NCAA**: 18/349, **CAA**: 3/256  **Egg/milk: NCAA**: 9/349, **CAA**: 4/256  **Antibiotic**: **NCAA**: 14/349, **CAA**: 8/256  **Total**: **NCAA**: 82/349, **CAA**: 20/256 | - | **IgE** **pollen**: <0.001  **Fur/mite**: 0.008  **Egg/milk**: 0.26  **Antibiotic**: 0.34  **Total**:  <0.001 |
| Kim et al. (2019) [20] | Korea | Retrospective cohort | 0 to > 85y | 58825/51325 | 110150 | 22030 | 88120 | **Appendectomy group:** 3358/22030 (15.2%) had asthma  OR: 1.18 (95% CI 1.13-1.23)  **Control group:** 11749/88120 (13.3%) had asthma | P<0.01 | - |
| Omling et al. (2021)* [23] | Sweden | Retrospective Cohort | 0-14y | 3658/2709 | 6367 | 6367  **NCAA**:5246, **CAA**:1121 | - | **IgE-mediated allergy: NCAA:** 1178/5246, **CAA:** 173/1121 | - | <0.001 |
| Carvalho et al. (2021) [22] | Portugal | Prospective case-control | >18y | 59/75 | 134 | 65  **NCAA**: 38, **GA**: 27 | 69  **IA**: 52  **NHF**: 17 | **Allergy: IA:** 8/51******, NHF:** 3/17**, NCAA:** 6/36******, GA:** 4/27 | 0.095 | - |
| Gudjonsdottir et al. (2022)* [25] | Sweden | Prospective cohort | <15y | 103/75 | 178 | 138 **NCAA**: 80**, CAA**: 58 | 40 | **Allergy: NCAA**: 15/80**, CAA**: 5/58 | - | 0.095***** |
| Zhang Z et al. (2023)* [27] | China | Retrospective cohort | 6.2-11.4y | 728/428 | 1156 | 1156  **CAA**: 517  **NCAA**: 639 | - | **IgE-mediated allergy: NCAA**: 105/639**, CAA**: 57/517 | - | 0.009 |
| Carvalho et al. (2024) [29] | Portugal | Prospective (case-control) | >18y | 32/28 | 60 | 46 **NCAA**: 22, **GA**: 24 | **IA**: 14 | **Allergy: IA**:1/13**, NCAA**: 2/20**, CAA**: 1/21 | - | 0.834 |

**Table 1. Summary of publications included assessing the role of allergy in acute appendicitis.**

**AA:** Acute appendicitis group; **CG**: Control group; **MPT**: Mixed Inhalant Prick Test; **FAT**: Food Allergy Test; **SPT**: Skin Prick Test; **SA**: Suppurative appendicitis; **PA**: Periappendicitis; **NCAA**: Non-complicated acute appendicitis; **CAA:** Complicated acute appendicitis; **GA**: gangrenous appendicitis; **IA**: Incidental appendectomy.

***1****: Mean (standard deviation)*

**Pediatric population; **: Units not reported; ***: The authors report only numerical values concerning the comparison between the different histological subtypes of acute appendicitis but not for the comparison between AA and CG. Similarly, there is a discordance between the results reported in the abstract (“A significant difference was determined between the patient and control groups in terms of skin prick positivity (p < 0.05)”) and the main text, where the opposite is stated ("No statistically significant difference was determined between the patient and control groups in terms of skin prick positivity (p > 0.05)"), ****: the sample size for this specific analysis was modified in the original paper (see table 1 from that paper), *****: all-groups comparison,******:* *It is inferred that the study is prospective, but the authors do not make this explicit.*
