## Supplementary material for "Role of type I hypersensitivity reaction in the development of overall and uncomplicated acute appendicitis: a systematic review and meta-analysis": Table 2

| **Author** | **Country** | **Study design** | **Age (Range)** | **Sex M/F** | **Total *N*** | ***N* in AA** | ***N* in CG** | **Type I Hypersensitivity reaction-related biomarkers assessment** | **P**  **(AA vs. CG)** | **P (NCAA vs. CAA)** |
| --- | --- | --- | --- | --- | --- | --- | --- | --- | --- | --- |
| Carvalho et al. (2019) [19] | Portugal | Prospective (case-control) | (4.9-53.1) | 24/17 | 41 | 33  **NCAA**: 20 **GA**: 13 | **NHF**: 8** | **IL-4 (ALF):**  **NCAA**: 158.1 (228), **CAA**: 109.2 (154.7),  **CG** : 23.5 (6.8) ^1, ***^  **IL-5 (ALF):**  **NCAA** 76.3 (106.5), **CAA**: 49.2 (81.6), **CG** : 9.9 (4) ^1, ***^  **IL-9 (ALF):**  **NCAA** 101.9 (141.7), **CAA**: 72.5 (110.3), **CG** : 17.9 (7) ^1, ***^ | **IL-4 (ALF):** 0.017****  **IL-5 (ALF):** 0.05****  **IL-9 (ALF):** 0.062**** | **IL4 (ALF):** 0.421  **IL-5 (ALF):** 0.382  **IL-9 (ALF):** 0.587 |
| Carvalho et al. (2021) [22] | Portugal | Prospective (case-control) | >18y | 59/75 | 134 | 65  **NCAA** : 38  **GA** : 27 | 69  **IA** : 52  **NHF** : 17 | **IgE (IHQ):**^*****^  **IA:** 17 (4.8-29), **NHF:** 26 (15-44), **NCAA:** 28 (15.8-61.8), **GA:** 22 (10.5-42) ^2^ | < 0.0001 (NCAA vs. IA) | NS |
| Carvalho et al. (2022) [24] | Portugal | Prospective case-control | >18y | 53/30 | 83 | **AA**: 65  **NCAA** : 38  **GA** : 27 | **NHF**: 18 | **IL-5 (SE):**  **NHF**: 14.38 (12-20.52), **NCAA**: 11.67 (9.43-18), **GA:** 13.05 (10.76-18.79) ^2, ***^  **NHF**: 15.6 (6.9), **NCAA**: 13.03 (6.6), **GA:** 14.2 (6.28) ^3, ***^  **IL-5 (ALF): NHF**: 8 (6.67-13.6), **NCAA**: 29.22 (9.06-115.16) **GA:** 8.67 (6.67-46.67) ^2, ***^ | **IL-5 (SE):** 0.239****  **IL-5 (ALF):** 0.056**** | - |
| Gudjonsdottir et al. (2022)* [25] | Sweden | Prospective cohort | <15 | 103/75 | 178 | 138 **NCAA**: 80 **CAA**: 58 | 40 | **IgE (SE, ng/mL):**  **NCAA**: 160.8 (72.6-530.5), **CAA**: 127.2 (64-404), **CG**: 121.6 (83.4-258.4) ^2^  **NCAA**: 254.6 (345.7), **CAA**: 198.4 (258.5), **CG**: 154.5 (134.6) ^3^  **IL-4 (SE):**  **NCAA**: 0.3 (0.2-0.6), **CAA**: 0.4 (0.2-0.7), **CG**: 0.4 (0.2-0.8) ^2, ***^  **NCAA**: 0.37 (0.3), **CAA**: 0.44 (0.38), **CG**: 0.47 (0.46) ^3, ***^  **IL-9 (SE):**  **NCAA**: 1.4 (0.8-2.1), **CAA**: 1.8 (1.1-3.2), **CG**: 1.9 (1-2.7) ^2, ***^  **NCAA**: 1.44 (0.98), **CAA**: 2.03 (1.60), **CG**: 1.87 (1.31) ^3, ***^  **IL-13 (SE):**  **NCAA**: 14.6 (10.2-24.3), **CAA**: 24.6 (12.9-58.5), **CG**: 11.3 (8.4-20.2) ^2, ***^  **NCAA**: 16.37 (10.64), **CAA**: 32 (34.67), **CG**: 13.3 (9.07) ^3, ***^ | **IgE (SE):** 0.326****  **IL-4 (SE):**  0.852****  **IL-9 (SE):**  0.088****  **IL-13 (SE):**  0.001**** | **IgE (SE):** 0.159  **IL-4 (SE):**  0.896  **IL-9 (SE):**  0.047  **IL-13 (SE):**  0.002 |
| Zhang T et al. (2023)* [28] | China | Prospective cohort | 2.5-15 | 79/61 | 140 | 140  NCAA: 97  GA: 43 | - | **IgE (SE, ng/mL): NCAA**: 154.29 (61.52), **GA**: 61.3 (30.33) ^1^  **IL-4 (SE): NCAA**: 0.14 (0.09), **GA**: 0.16 (0.08) ^1, ***^  **IL-5 (SE): NCAA**: 6.27 (3.26), **GA**: 6.58 (3.44) ^1, ***^  **IL-6 (SE): NCAA**: 5.64 (2.25-10.59), **GA**: 16.06 (12.4-70) ^2, ***^  **NCAA**: 6.16 (6.28), **GA**: 32.82 (44.2) ^3, ***^  **IL-9 (SE): NCAA**: 0.93 (0.47), **GA**: 1.2 (0.93) ^1, ***^  **IL-13 (SE):NCAA**: 7.54 (3.95), **GA**: 18.5 (8.57) ^1, ***^ | - | **IgE (SE):** <0.01  **IL-4 (SE):** 0.201  **IL-5 (SE):** 0.613  **IL-6 (SE):** <0.001  **IL-9 (SE):** 0.029  **IL-13 (SE):** <0.01 |
| Carvalho et al. (2024) [29] | Portugal | Prospective (case-control) | >18y | 32/28 | 60 | 46 **NCAA**: 22 **GA**: 24 | **IA**: 14^X^ | **Histamine (ALF):**  **NCAA**: 1.2 (0.6-2), **CAA**: 1.2 (0.6-1.9), **CG**: 1.8 (0.4-1.9) ^2^  **Serotonin (ALF):**  **NCAA**: 214.25 (80.5-633.12), **CAA**: 87 (22.5-589.95), **CG**: 221.25 (89.5-229.5) ^2^  **Tryptase (ALF):**  **NCAA**: 53.95 (38.77-135.18), **CAA**: 92.58 (50.16-134.26), **CG**: 8.25 (5.6-11.5) ^2^ | **Histamine (ALF):**  0.948****  **Serotonin (ALF):**  0.290****  **Tryptase (ALF):**  0.001**** | - |

**Table 2. Summary of publications included assessing the role of type I Hypersensitivity-related biomarkers in acute appendicitis.**

**AA:** Acute appendicitis group; **CG**: Control group; **GA**: Gangrenous appendicitis; **ALF**: Appendicular lavage fluid; **IA**: Incidental appendectomy; **NHF**: Normal histologic findings, **IHQ**: Immunohistochemical staining; **NS**: Non-significant; **SE**: Serum; **NCAA**: Non-complicated acute appendicitis; **CAA:** Complicated acute appendicitis

***1****: Mean (standard deviation),* ***2****: Median (interquartile range),* ***3****: Mean (standard deviation) estimation from median (interquartile range)*

**: Pediatric population; **: CG defined as a non-pathological appendix (histological evaluation), ***: pg/mL, ****: 3-groups comparison, *****: number of IgE positive staining cells (mast cells).* ***X****: Patients diagnosed with colorectal carcinoma.*
