## Supplementary material for "Role of type I hypersensitivity reaction in the development of overall and uncomplicated acute appendicitis: a systematic review and meta-analysis": Table 3

| **Author** | **Country** | **Study design** | **Age (Range)** | **Sex M/F** | **Total *N*** | ***N* in AA** | ***N* in CG** | **Eosinophils/mast cells assessment** | **P**  **(AA vs CG)** | **P (NCAA vs CAA)** |
| --- | --- | --- | --- | --- | --- | --- | --- | --- | --- | --- |
| Harlak et al. (2008) [12] | Turkey | Prospective case-control | 14-57y | 171/40 | 211 | 111 | **CG**: 100 | **EIR grade 2**:  MPT(+): 2/33, MPT(-): 4/78  **EIR grade 3**:  MPT(+): 3/33, MPT(-): 4/78  **TEC (PB) (/mm^3^)**:  **AA**: 100 (71.7) ^1^,  **NHF**: 86.5 (74.1)^1^  **TEC (PB) (x10^9^/L):**  **AA**: 0.1 (0.07) ^1^, **CG**: 0.08 (0.07)^1, XX^ | **EIR**: 0.664  **TEC**: 0.18 | - |
| Santhosh et al. (2008) [13] | India | Prospective case-control | - | - | 32 | 18 | 14  **NHF**: 6 **ABA**: 3  **HV**: 5 | **SECP (****ng/mL):**  **AA:** 31.7 (9.7-175) ^2^  **NHF**: 23.6 (8.8-30.6) ^2^  **ABA**: 17.3 (11.4-43.9) ^2^  **HV**: 16 (6.1-18.8) ^2^  **MEC (n/hpf)**  **AA:** 16.6/hpf  **NHF:** 1.2/hpf | **SECP**  **AA vs HV:** 0.007  **MEC**  **AA vs NHF:**  0.0003 | - |
| Kolur et al. (2014) [14] | India | Retrospective and prospective case-control | 6-75y | 385/392 | 777 | **AA**: 340  **SA**: 18  **EA**: 23  **RA**: 356 | **NHF**: 40 | **MEC (total mean eosinophilic count/mm^2^)**  Mucosa: **NHF**: 28.4 (12.5), **AA**: 23.7 (10.7), **SA**: 25.9 (11.8), **EA**: 69 (30.2), **RA**: 34.2 (18.6)^1^  Submucosa: **NHF**: 26.2 (11.5) **AA**: 24.5 (11.3), **SA**: 26.1 (10.3), **EA**: 48 (29.1), **RA**: 30 (18.2)^1^  M. propria: **NHF**: 21.2 (10.3) **AA**: 22.4 (9.7), **SA**: 18.5 (6.6), **EA**: 44.6 (23.9), **RA**: 28.2 (16.8)^1^  **MCC (total mean mast cell count /mm^2^)**  Mucosa: **NHF**: 7.2 (3.7), **AA**: 8.8 (4.2), **SA**: 8.3 (3.4), **EA**: 19.5 (11.5), **RA**: 11.3 (6.1)^1^  Submucosa: **NHF**: 8.15 (4.7), **AA**: 8.48 (4.48), **SA**: 8.27 (5.26), **EA**: 16.6 (12.8), **RA**: 10.2 (5.8)^1^  M. propria: **NHF**: 9.38 (4.9), **AA**: 8.16 (3.8), **SA**: 9.05 (5), **EA**: 16.3 (9.3), **RA**: 9.99 (5.6)^1^ | **-** | - |
| Karakuş et al. (2015)* [15] | Turkey | Not specified | **AA**: 9.9 ^6^  **CG**: 6.7 ^6^ | - | 40 | 30 | **NHF**: 10 | **Cajal Cells density mean values: CG**: 8.3, **AA**: 4.93  **Eosinophils density mean values: CG**: 4.7, **AA**: 21.23  **Mast cells density mean values**:  **CG**: 14.7, **AA**: 22.53 | **Cajal cells:** <0.05  **Eosinophils:** <0.05  **Mast cells:** <0.05 |  |
| Cevizci et al. (2016)* [16] | Turkey | Prospective case-control****** | **AA**: 10.1 (2.9)y ^1^  **CG**: 9.8 (2.6)y ^1^ | 140/100 | 240 | 116 | 124 | **TEC (PB):**  **AA**: 1.31 (0.6), **CG**: 1.18 (0.4)^1, **^  **TEC (PB) (x10^9^/L):**  **AA**: 0.131 (0.06), **CG**: 0.118 (0.04)^1, **^ | **TEC**: >0.05 | >0.05 |
| Minderjahn et al. (2018)* [17] | China | Retrospective cohort | 7-14y | 390/257 | 647 | **NCAA:** 395 **CAA:** 252 | - | **TEC (PB) (x10^9^/L):**  **NCAA :** 0.11 (0.19)^,^ **CAA**: 0.046 (0.104)^1^ | - | <0.0001 |
| Mohan et al. (2020) [21] | India | Prospective case-control | - | - | 25 | **AA**:15 | **NHF**: 10 | **MEC (eosinophils in muscle layer/mm^2^):**  **AA**: 72.9 (33.8-112)^3^  **NHF**: 2.1 (0.4-4.7)^3^  **MEC (eosinophils in muscle layer per whole cross-sectional area):**  **AA**: 1450.7 (464.2–2437.2)^3^  **NHF**: 13.6 (3.9-31.0)^3^  **MCC (degranulated mast cells in submucosa per whole cross-sectional area):**  **AA**: 1136.4 (641.6-1631.2)^3^  **NHF**: 285.3 (28-598.6)^3^ | **MEC (muscle layer/mm^2^)**: 0.005  **MEC (muscle layer, whole cross-sectional area):** 0.02  **MCC (submucosa, whole cross-sectional area):** 0.01 | - |
| Carvalho et al. (2022) [24] | Portugal | Prospective case-control | >18y | 53/30 | 83 | **AA**: 65  **NCAA**:38  **GA**: 27 | **NHF**: 18 | **TEC (PB) (x10^9^/L):**  **NHF**: 0.13 (0.004-0.22), **NCAA**: 0.08 (0.03-0.15), **GA**: 0.03 (0.02-0.07)^4^  **NHF**: 0.118 (0.173), **NCAA**: 0.086 (0.09), **GA**: 0.04 (0.039)^5^  **EIR (AW) absolute number, 10 hpf**  **NHF**: 47.5 (24-62), **NCAA**: 1 (37.5-95.5) X, **GA**: 21 (9-34)^4^ | **TEC**: 0.004******  **EIR (AW):** <0.001****** | - |
| Carvalho et al. (2023) [26] | Portugal | Prospective case-control | >18y | 32/28 | 60 | **AA**: 46  **NCAA**: 22  **GA**: 24 | **NHF**: 14 | **TEC (PB) (x10^9^/L):**  **NHF**: 0.04 (0.01-0.21), **NCAA**: 0.14 (0.04-0.26), **GA**: 0.02 (0.00-0.07)^4^  **NHF**: 0.086 (0.164), **NCAA**: 0.147 (0.174), **GA**: 0.03 (0.055)^5^  **EDN (ALF, ng/mL):**  **NHF**: 4.93 (3.2-5.43) **NCAA**: 5.23 (4.88-6.9), **GA**: 5.02 (3.55-5.65)^4^  **ECP (ALF, ng/mL):**  **NHF**: 3.35 (2.6-6.1), **NCAA**: 38.85 (26.5-51.77), **GA**: 51.55 (39.55-70.09)^4^  **EP (ALF, ng/mL):**  **NHF**: 45.55 (39.9-74), **NCAA**: 240.28 (191.2-341.3), **GA**: 302.5 (227.7-535.85)^4^  **EDN (SE, ng/mL):**  **NHF**: 4.58 (1.63), **NCAA**: 3.48 (1.13), **GA**: 4.21 (1.54)^1^  **SECP (ng/mL):**  **NHF**: 7.56 (4.76-11.13), **NCAA**: 39 (21.3-56.9), **GA**: 51.3 (20.25-62.59)^4^  **EP (SE, ng/mL):**  **NHF**: 38.2 (30.2-59.1), **NCAA**: 158.4 (111.09-222.1), **GA**: 235.27 (192.33-262.51)^4^ | **TEC (SE)**: 0.052******  **EDN (ALF):** 0.256****  **ECP (ALF):** <0.001****  **EP (ALF): <** 0.001****  **EDN (SE):** 0.067****  **SECP (SE):** <0.001****  **EP (SE): <** 0.001**** | - |

**Table 3. Summary of publications included assessing the role of eosinophil and mast cells in acute appendicitis.**

**AA:** Acute appendicitis group; **CG**: Control group; **GA**: Gangrenous appendicitis; **IA**: Incidental appendectomy; **NHF**: Normal histologic findings; **IHQ**: Immunohistochemical staining; **ALF**: Appendicular lavage fluid, **NS**: Non-significant; **SE**: Serum; **ABA**: Acute bronchial asthma; **HV**: Healthy volunteers; **SECP**: Serum eosinophil cationic protein; **EDN**: Eosinophil-derived neurotoxin; **EP**: Eosinophil peroxidase; **MEC**: Mean eosinophilic count; **EIR**: Eosinophilic infiltration rate; **AW**: Appendiceal wall; **TEC**: Total eosinophil count; **hpf**: High power field; **PB**: Peripheral blood; **NCAA**: Non-complicated acute appendicitis; **CAA:** Complicated acute appendicitis; **EA**: Acute eosinophilic appendicitis; **RA**: Recurrent appendicitis; **MCC:** Mast cell count

***1****: Mean (standard deviation);* ***2****: Median (range);* ***3****: Mean (95%CI);* ***4****: Median (interquartile range)* ***5****: Mean (standard deviation) estimation from median (interquartile range)* ***6****: Mean*

**Pediatric population; **: Units not reported, ***: pg/mL, ****: 3-groups comparison, *****: Number of IgE positive staining cells (mastocytes), ******: It is inferred that the study is prospective, but the authors do not make this explicit.* ***X****: We believe this number, “1(37.5-95.5),” is a mistake since it is not mathematically possible.* ***XX****: Conversion from /mm3 to /1x10^9/L.*
