## Supplementary File 1 for "Role of type I hypersensitivity reaction in the development of overall and uncomplicated acute appendicitis: a systematic review and meta-analysis"

**Exclusion criteria**

-Case reports.

-Duplicate or overlapping studies.

-Reviews, systematic reviews, consensus guidelines.

-Languages other than English or Spanish.

-Studies with no population of interest.

-Studies conducted in immunocompromised patients.

-Studies conducted in patients with a previously diagnosed immune disorder.

-History of using antihistaminic or antiallergenic drugs in the previous week.

-Studies conducted in patients with metastatic neoplastic disease.

**Inclusion criteria**

-Pilot prospective or retrospective studies evaluating the role of immunoallergic mediation in the development of acute appendicitis.

-Pilot prospective or retrospective observational original clinical studies measuring the diagnostic performance of allergy biomarkers compared to the reference standards for the diagnosis of appendicitis and for the discrimination between complicated and uncomplicated appendicitis.

-Diagnostic validation original studies measuring the diagnostic performance of allergy biomarkers. compared to the reference standards for the diagnosis of appendicitis and for the discrimination between complicated and uncomplicated appendicitis.

**Supplementary file 1. Inclusion and exclusion criteria**
